# A deep learning algorithm based on fundus photographs to measure retinal vascular parameters and their additional value beyond the CAIDE risk score for predicting 14-year dementia risk

**DOI:** 10.1101/2025.08.26.25334432

**Authors:** Darui Gao, Yanyu Zhang, Jianhao Xiong, Sijin Zhou, Yanjun Ma, Yang Pan, Zongyuan Ge, Xiangang Chang, Hongyu Wang, Bin Lv, Fanfan Zheng, Wuxiang Xie

## Abstract

**Background:** Retinal photography is a valuable non-invasive tool for assessing the nature of vessel changes. It is of interest whether retinal vascular parameters can improve the ability to predict dementia risk of the widely used Cardiovascular Risk Factors, Aging, and Incidence of Dementia (CAIDE) model.

**Methods:** A fully automated artificial intelligence algorithm providing measures of seven meaningful parameters of the retinal vascular was developed and evaluated. Applying this algorithm to analyze the fundus images collected by the UK Biobank (UKB) study and the Beijing Research on Ageing and Vessel (BRAVE) study, we further explore the associations between retinal vascular parameters with arteriosclerosis and cognitive function across two countries. Finally, we fitted a published algorithm-estimated CAIDE model based on fundus images developed by our research group to the UKB study and evaluated the additional value of retinal vascular parameters beyond the algorithm-estimated CAIDE model for predicting 14-year dementia risk.

**Findings:** In the UKB cohort (n=35,838), the algorithm-estimated CAIDE model achieved an area under the curve (AUC) of 0.697 (95% confidence interval [CI]: 0.678-0.717) for 14-year all-cause dementia, which was comparable to that of the actual CAIDE model (AUC=0.683, 95% confidence interval: 0.663-0.703, p=0.272). Furthermore, adding retinal vascular parameters to the algorithm-estimated CAIDE model increased the AUC statistically significantly from 0.692 to 0.711 (p<0.001) in the derivation set (n=38,384), and from 0.682 to 0.706 (p=0.018) in the internal validation (n=9594) for all-cause dementia.

**Interpretation:** The integration of automatically extracted retinal vascular parameters into the algorithm-estimated CAIDE model improves the predictive ability of 14-year dementia risk. Compared to the original CAIDE model, the retinal vascular parameters-enhanced algorithm-estimated CAIDE model may provide a more accurate dementia risk assessment with just a single fundus photograph.

**Funding:** The Capital’s Funds for Health Improvement and Research, and the National Natural Science Foundation of China.

## Introduction

On a global scale, it is estimated that the number of individuals with dementia will increase from 57.4 million cases in 2019 to 152.8 million cases by 2050, driven by population growth and aging.^1^ However, there is currently no effective and widely available treatment. Accumulating evidence suggests that up to 45% of dementia cases could be prevented or delayed by modifying 14 risk factors associated with daily living.^2^ Therefore, the early identification of high-risk individuals with modifiable risk factors can enhance their awareness of the risks and enable them to effectively decrease their risk of dementia through lifestyle management.

To date, various dementia risk prediction scores have been developed according to varying requirements and settings. However, most of these lack high-quality external validation and practical experience in real-world applications.^3–5^ The Cardiovascular Risk Factors, Aging, and Incidence of Dementia (CAIDE) dementia risk score is a well-established, validated model widely used to predict the 20-year risk of developing dementia and adopted in the Finnish Geriatric Intervention Study (FINGER) to select eligible at-risk participants.^6–8^ The calculation of the CAIDE score requires information obtained through processes such as questionnaires, physical examinations, and fasting blood draws. These procedures are time-consuming and invasive, increasing the workload for healthcare professionals. Our research team has pioneered the use of retinal images from 292,554 participants to train and validate a deep learning algorithm aimed at estimating the CAIDE dementia risk score. This approach provides an efficient, convenient, and non-invasive tool for screening dementia risk in large populations.^9^ However, the algorithm-estimated CAIDE score has not yet been validated in longitudinal studies with dementia events.

Furthermore, although the developers of the original CAIDE model emphasized the close association between vascular health and dementia, incorporating components such as blood pressure and cholesterol—strongly correlated with vascular health—into the model, it did not include indicators that directly reflect an individual’s vascular health status.^7^ With recent advances in artificial intelligence (AI) recognition and deep learning capabilities, the accurate and automatic segmentation of retinal vessel and measurement of retinal vascular parameters eventually become key areas of interest.^10,11^ Retinal vessel morphometry assessment provides insight into the nature of vessel changes, and the pathological relationship between retinal microvascular changes and systemic vascular abnormalities has been well recognized.^12–14^

Therefore, we fitted the algorithm-estimated CAIDE model to a large-scale prospective cohort study and aimed to assess the potential of improving its ability to predict dementia risk by including retinal vascular parameters. To achieve this objective, we developed a deep learning algorithm for the fully automated extraction of retinal vascular features and investigated the associations of these parameters with biomarkers of arteriosclerosis and cognitive function across two countries.

## Methods

### Study design, datasets, and participants

We conducted a multistage population-based study in five parts using prospective and cross-sectional datasets. The detailed description of the study design is provided in the **appendix (pp 4-6)** and **Figure 1**. The detailed flowcharts outlining the inclusion and exclusion criteria for participants at each stage are presented in the **appendix (pp** 7-10).

**Figure 1.**
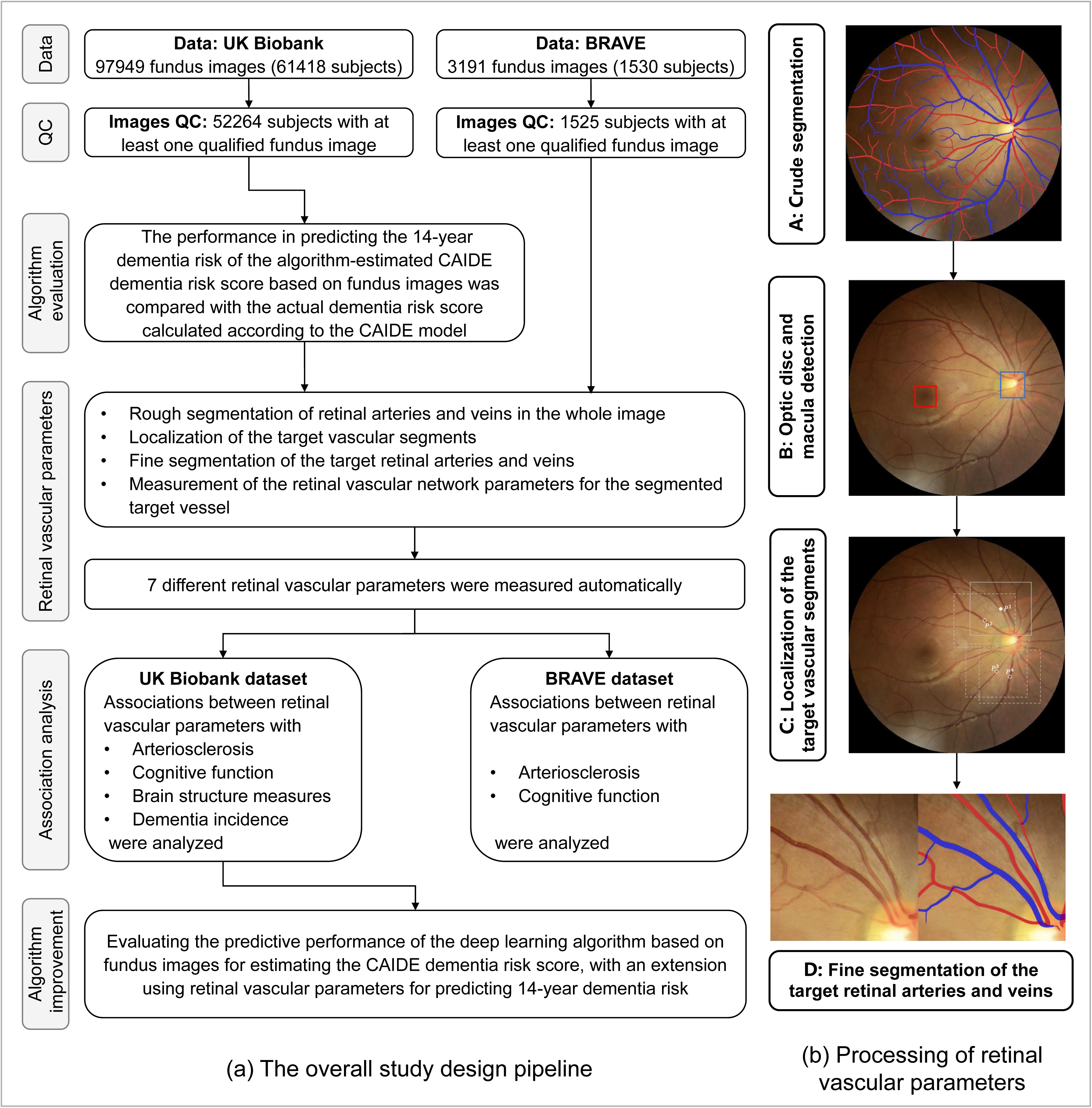
The overall study design pipeline and processing of retinal vascular parameters. (a) The overall study design pipeline; (b) Processing of retinal vascular parameters; A: Visualization of retinal arteriovenous crude segmentation. Red represents arteries, blue represents veins. B: Visualization of optic disc and macula detection; The red box represents the macular area, and the blue box represents the optic disc. C: Localization of the target vascular segments; D: Fine segmentation of the target retinal arteries and veins. Red represents arteries, blue represents veins.

Firstly, we used a published deep learning algorithm^9^ developed by our research group for estimating the CAIDE dementia risk score via fundus images to obtain an algorithm-estimated score for each participant with qualified fundus photographs in the UK Biobank (UKB) study. This involved analyzing 97,949 fundus images from 61,418 subjects, with ethical approval obtained from the North West Multi-Centre Research Ethics Committee (Research Ethics Committee reference: 16/NW/0274) and informed consent obtained for all participants.^15,16^ To compare the predictive performance of the algorithm-estimated CAIDE score with the actual CAIDE model-computed score, a combination of variables derived from the questionnaire and blood samples was also collected. According to the inclusion and exclusion criteria outlined in the **appendix (pp 7)**, a total of 35,838 participants were included in the analysis for this stage. The demographic characteristics of these participants are summarized in the **appendix (pp 11)**.

Secondly, we developed and evaluated an automatic quantitative analysis algorithm for measuring retinal vascular parameters. The training set and evaluation set comprised 860 fundus images sourced from our internal Chinese medical check-up database and the publicly available AV-DRIVE database.^17^

Thirdly, the algorithm mentioned above was applied to the fundus images collected by the UKB study and the Beijing Research on Ageing and Vessel (BRAVE) study for the segmentation and quantification of retinal microvasculature. The BRAVE study is a community-based, prospective cohort study investigating the contributions of vascular conditions to cognitive impairment and dementia.^18^ The study was approved by the Institutional Review Board of Peking University Health Science Center (IRB0001052-19060) and Fuwai Hospital (IRB2012-BG-006), and all participants gave their written informed consent according to the Declaration of Helsinki. A total of 1554 participants were initially enrolled and underwent baseline evaluation. Of these, 3191 fundus images were collected from 1530 subjects during baseline assessments. The demographic characteristics of 1506 participants who had complete data on retinal vascular parameters and the traditional CAIDE model are summarized in the **appendix (pp 11)**.

Fourthly, we investigated the associations between retinal vascular parameters with arteriosclerosis and cognitive performance in the UKB (n=38,630 and 37,762, respectively) and BRAVE studies (n=1320 and 1500, respectively), respectively. In the UKB study, we conducted a further analysis of the relationship between retinal vascular parameters and brain structural measures and dementia incidence (n=3699 and 38,863, respectively).

Finally, the UKB dataset was randomly split into a derivation (80%, n=38,384) and a validation set (20%, n=9594) to assess whether 14-year dementia risk prediction with the estimated CAIDE dementia risk score via fundus images can be improved by incorporating retinal vascular parameters.

### Processing of the estimated CAIDE dementia risk score derived from fundus images

Retinal images were processed using a deep learning algorithm developed by our research group for estimating the CAIDE dementia risk score.^9^ In brief, this CAIDE estimated algorithm was trained and tested using the InceptionResNetV2 architecture on the platform Keras v2.2.2 and the Python scikit-learn package 0.22.2. The R² between the estimated and actual CAIDE dementia risk scores was 0.80 for the internal validation dataset and 0.58 for the external validation dataset.

### Variables of the CAIDE model

The CAIDE model is a validated dementia risk prediction tool developed from a Finnish population-based cohort study involving 1,409 participants.^7^ The CAIDE model includes age, education, sex, systolic blood pressure, body mass index (BMI), total cholesterol, and physical activity. All variables included in the CAIDE model were available in both the UKB and BRAVE studies. To mimic clinical practice, we did not impute missing data. According to previous studies,^9,19–21^ a comprehensive description of the variables in the CAIDE model is provided in the **appendix (pp** 12-15).

### Processing of retinal vascular parameters

In this study, we developed an AI algorithm to process retinal images. The detailed methods of algorithm development and evaluation are described in the **appendix (pp 16-19)**. In brief, this algorithm achieves the fully automated measurement of retinal vascular parameters through four steps, including (1) rough segmentation of retinal arteries and veins in the whole image; (2) localization of the target vascular segments; (3) fine segmentation of the target retinal arteries and veins; and (4) measurement of the retinal vascular network parameters for the segmented target vessel.

We evaluated our proposed algorithm with two different tasks. The first task was conducted on our internal image dataset. We used mean Intersection over Union (mIoU) as the metric to evaluate the performance of segmentation.^22,23^ In addition, we evaluated the proposed algorithm on the AV-DRIVE database to verify the robustness. In this external evaluation, we adopted four metrics for the evaluation of vessel segmentation: the average accuracy (Acc), sensitivity (Sen), specificity (Spe), and area under the curve (AUC).^24^ A/V classification performance was evaluated using pixel-wise Acc, Sen, and Spe for the ground-truth artery vein pixels.^24^ By taking arteries as positives and veins as negatives, Sen reflects the proposed algorithm’s capability of detecting arteries and Spe for veins.

By utilizing this algorithm, we achieved a fully automated quantitative measurement of seven meaningful parameters of the retinal vascular, including central retinal arterial equivalent (CRAE), central retinal venular equivalent (CRVE), arteriovenous ratio (AVR), fractal dimension artery (FDa), fractal dimension vein (FDv), tortuosity of artery, and tortuosity of vein. Detailed descriptions of these parameters are provided in the **appendix (pp 20)**.

### Dementia incidence

Incident dementia and corresponding date in the UKB study were ascertained using diagnoses obtained from the Hospital Episode Statistics for England, the Patient Episode Database for Wales, and death register data from National Health Service Digital. A detailed definition can be found on the UKB website (https://biobank.ndph.ox.ac.uk/showcase/refer.cgi?id=460) and in the **appendix (pp 21-24)**. According to a validation study, the algorithms developed for ascertaining dementia cases, with data sources of hospital records and death registers, reached positive predictive values of 84.5% for all-cause dementia, 70.8% for Alzheimer’s disease (AD), and 33.3% for vascular dementia (VD).^25^ We excluded participants with a diagnosis of dementia before cohort entry, and followed up for the participants from baseline until the first incidence of dementia, death, loss to follow-up, or the end of the study (June 30, 2024), whichever came first.

### Brain MRI measures

Brain MRI imaging was conducted as an extension of the UKB study. Further details regarding image acquisition and processing are available in the protocol.^26^ We used data from the baseline measurement since 2014. Volumes of specific brain structures, including peripheral cortical gray matter, gray matter, white matter, and total brain, were made available by the UKB Imaging team as image-derived phenotypes. All volume measures were adjusted for head size using a SIENAX-style analysis.^27^

### Arteriosclerosis measurement

Arteriosclerosis assessment was conducted using different metrics across the two cohorts. In the UKB study, arterial stiffness was measured noninvasively through pulse waveforms collected from the finger using an infra-red sensor (Pulse Trace PCA2, CareFusion). The Arterial Stiffness Index (ASI) in meters per second was calculated by dividing participants’ height (in meters) by the duration between the peaks of the pulse waveform.^14^

In the BRAVE study, brachial ankle pulse wave velocity (ba-PWV) was assessed using a non-invasive arteriosclerosis measuring device (Vascular Inspecting Apparatus MB3000, M&B Electronic Instruments, China).^18^ The mean value of ba-PWV from the left and right sides was used in the analyses. Coronary artery calcification (CAC) was measured using a multi-slice CT scan (Brilliance iCT, Philips Healthcare, Cleveland, OH, USA). The CAC score (CACS) was automatically calculated by manually adding all detectable calcification lesions.^28^ The detailed methods of assessment of ba-PWV and CACS were provided in the **appendix (pp 25)**.

### Cognitive measurements

In the UKB study, participants completed several brief cognitive tests via a touchscreen interface at the baseline assessment. In this study, we selected three cognitive tests to assess the cognitive function of the participants: reaction time (in milliseconds [ms]), fluid intelligence (correct responses), and prospective memory (dichotomous, correct first time). Detailed descriptions regarding these cognitive tests have been published elsewhere.^29^ Higher scores on the reaction time test indicate poorer cognitive performance, whereas higher scores on the fluid intelligence test reflect better cognitive performance. Additionally, a value of 1 on the prospective memory test also suggests better cognitive performance.

In the BRAVE study, the cognitive measurement was the Chinese version of the Montreal Cognitive Assessment Basic (MoCA-B), a sensitive and validated cognitive test battery to comprehensively assess nine cognitive domains.^30^ In addition, we also supplemented three tests to assess specific cognitive domains further (verbal memory, semantic fluency, and executive function); the detailed methods were provided in the **appendix (pp 26-27)**. All cognitive scores in the BRAVE were converted to standardized Z scores by subtracting the mean scores and dividing by the standard deviation (SD) to facilitate comparisons across tests, where a higher score indicates better cognitive performance.

### Statistical analysis

The results were presented using percentages for categorical variables and means ± SD for continuous variables. To evaluate the performance of each model to predict the 14-year risk of dementia, we further excluded participants who did not develop dementia and had less than 14 years of follow-up, and each participant was followed up to 14 years from the date of baseline assessment. The discriminative ability of models for 14-year dementia risk was determined by the area under the receiver operating characteristic (ROC) curve and 95% confidence intervals (CIs). DeLong’s test was performed for the statistical comparison of AUCs. Linear regression and logistic regression models, adjusted for the actual CAIDE score, were employed to examine the associations between retinal vascular parameters and arteriosclerosis, cognitive performance, and brain structural measures. Additionally, Cox proportional hazards regression models, also adjusted for the actual CAIDE score, were utilized to investigate the associations between retinal vascular parameters and the incidence of all-cause dementia, AD, and VD. Improvement in risk prediction with the addition of retinal vascular parameters to the algorithm-estimated CAIDE model via fundus images was assessed through the AUC.

All analyses were performed using SAS version 9.4 and R version 4.5.0. Statistical significance was defined as p value < 0.05; all tests were 2-tailed.

## Results

### Prediction performance comparison between the algorithm-estimated CAIDE model via fundus images and the actual CAIDE model

**Table 1** demonstrates the predictive performance of the algorithm-estimated CAIDE model for 14-year dementia risk via fundus images and provides a comparison with the actual CAIDE model. In the overall population of UKB study (n=35,838), the algorithm-estimated CAIDE model achieved AUCs of 0.697 (95% CI: 0.678-0.717), 0.702 (0.673-0.731), and 0.707 (0.658-0.756) for all-cause dementia, AD, and VD, respectively, which were comparable to those of the actual CAIDE model (all DeLong’s test p>0.05). Similar nonsignificant differences were observed in both women and men (all DeLong’s test p>0.05, **Table 1**).

**Table 1.** The predictive performance of the algorithm-estimated CAIDE model for 14-year dementia risk via fundus images and compared to the actual CAIDE model.

| **Metrics** | **Overall** | **Female** | **Male** |
| --- | --- | --- | --- |
| **All-cause dementia** |  |  |  |
| Events/Total | 509/35838 | 236/19393 | 273/16445 |
| AUC (Actual CAIDE model) | 0.683 (0.663-0.703) | 0.690 (0.661-0.720) | 0.667 (0.639-0.695) |
| AUC (Algorithm-estimated CAIDE model) | 0.697 (0.678-0.717) | 0.689 (0.661-0.718) | 0.695 (0.668-0.723) |
| DeLong's test p value for AUC comparisons | 0.272 | 0.953 | 0.115 |
| **Alzheimer's disease** |  |  |  |
| Events/Total | 236/35838 | 117/19393 | 119/16445 |
| AUC (Actual CAIDE model) | 0.679 (0.650-0.707) | 0.680 (0.640-0.721) | 0.674 (0.635-0.713) |
| AUC (Algorithm-estimated CAIDE model) | 0.702 (0.673-0.731) | 0.707 (0.666-0.747) | 0.692 (0.650-0.735) |
| DeLong's test p value for AUC comparisons | 0.196 | 0.324 | 0.453 |
| **Vascular dementia** |  |  |  |
| Events/Total | 79/35838 | 35/19393 | 44/16445 |
| AUC (Actual CAIDE model) | 0.751 (0.710-0.792) | 0.759 (0.694-0.824) | 0.735 (0.681-0.790) |
| AUC (Algorithm-estimated CAIDE model) | 0.707 (0.658-0.756) | 0.706 (0.638-0.775) | 0.700 (0.629-0.770) |
| DeLong's test p value for AUC comparisons | 0.138 | 0.221 | 0.417 |

### Deep learning for automatic vessel segmentation and A/V classification

The visualization results of automatic vessel segmentation and A/V classification are shown in **Figure 1 (b)**. The results of the mIoU of human-human and human-algorithm are reported in the **appendix (pp 28)**. MIoU of human-human arteries and veins were 0.472 and 0.524, respectively. While the mIoU of the human-algorithm for arteries was 0.451, and for veins was 0.524. The performance comparison between existing methods and our proposed algorithm on the AV-DRIVE database is presented in the **appendix (pp 29-30)**. In the comparison of vessel segmentation performance, our proposed algorithm achieves 91.76% Sen and 98.38% AUC, which is superior to the existing methods.^24,31–33^ However, compared to existing research,^34–37^ our proposed algorithm performs slightly worse in A/V classification, with Acc, Spe, and Sen of 90.1%, 91.8%, and 89.7%, respectively.

### Association of retinal vascular parameters with arteriosclerosis and cognitive performance in the UKB and BRAVE study

We first examined the associations between retinal vascular parameters with arteriosclerosis, as outlined in **Table 2**. The FDa showed inverse associations with ASI, CACS, and ba-PWV after adjustment for the actual CAIDE score (β = -0.091, -53.458, and -0.490, respectively, all p<0.001). The inverse associations between FDv with CACS and ba-PWV were also observed, with β of -49.950 and -0.325, respectively (both p<0.001). Among the retinal vessel diameter-related parameters, increased AVR and CRAE were both associated with a decrease in ASI (β=-0.133 and -0.076, respectively, both p<0.001) and CACS (β=-17.406 and -23.342, respectively, both p<0.05). In contrast, a 1 SD higher CRVE was only significantly associated with a 0.070 increase in ASI (p<0.001).

**Table 2.** Association of retinal vascular parameters with arteriosclerosis in the UKB and BRAVE study.

| Retinal vascular parameters | UKB (n=38630) |  | BRAVE (n=1320) |  |  |  |
| --- | --- | --- | --- | --- | --- | --- |
|  | Pulse wave Arterial Stiffness index |  | Coronary artery calcification scores |  | ba-PWV |  |
| | $\beta$ (95% CI) <sup>a</sup> | p value | $\beta$ (95% CI) <sup>a</sup> | p value | $\beta$ (95% CI) <sup>a</sup> | p value |
| AVR, per SD | -0.133 (-0.165, -0.100) | **<0.001** | -17.406 (-34.664, -0.148) | **0.048** | -0.103 (-0.236, 0.031) | 0.131 |
| CRAE, per SD | -0.076 (-0.108, -0.043) | **<0.001** | -23.342 (-40.57, -6.114) | **0.008** | -0.045 (-0.178, 0.089) | 0.510 |
| CRVE, per SD | 0.070 (0.038, 0.102) | **<0.001** | -5.644 (-22.960, 11.673) | 0.523 | 0.087 (-0.047, 0.221) | 0.201 |
| FDa, per SD | -0.091 (-0.125, -0.058) | **<0.001** | -53.458 (-71.478, -35.439) | **<0.001** | -0.490 (-0.628, -0.351) | **<0.001** |
| FDv, per SD | -0.017 (-0.049, 0.016) | 0.326 | -49.950 (-67.465, -32.435) | **<0.001** | -0.325 (-0.461, -0.189) | **<0.001** |
| Tortuosity of artery, per SD | -0.004 (-0.036, 0.029) | 0.822 | -19.037 (-36.343, -1.731) | 0.031 | -0.102 (-0.235, 0.032) | 0.137 |
| Tortuosity of vein, per SD | 0.032 (0.000, 0.064) | 0.053 | -2.971 (-20.282, 14.340) | 0.737 | 0.035 (-0.099, 0.169) | 0.606 |

<sup>a</sup> Adjusted for the actual CAIDE score.
Abbreviations: AVR: Arteriovenous ratio; CRAE: Central retinal arterial equivalent; CRVE: Central retinal venular equivalent; FDa: Fractal dimension artery; FDv: Fractal dimension vein.

**Table 3** presents associations of retinal vascular parameters with cognitive performance. The results indicated that higher FDa and FDv were significantly associated with improved cognitive performance across various domains after adjustment for the actual CAIDE score, including reaction time, prospective memory, verbal memory, executive function, and MoCA score (all p<0.05). Increased tortuosity of artery was also associated positively with reaction time, prospective memory, and verbal memory (β=-3.434 and 0.056, and OR=1.096, respectively, all p<0.001).

**Table 3.**
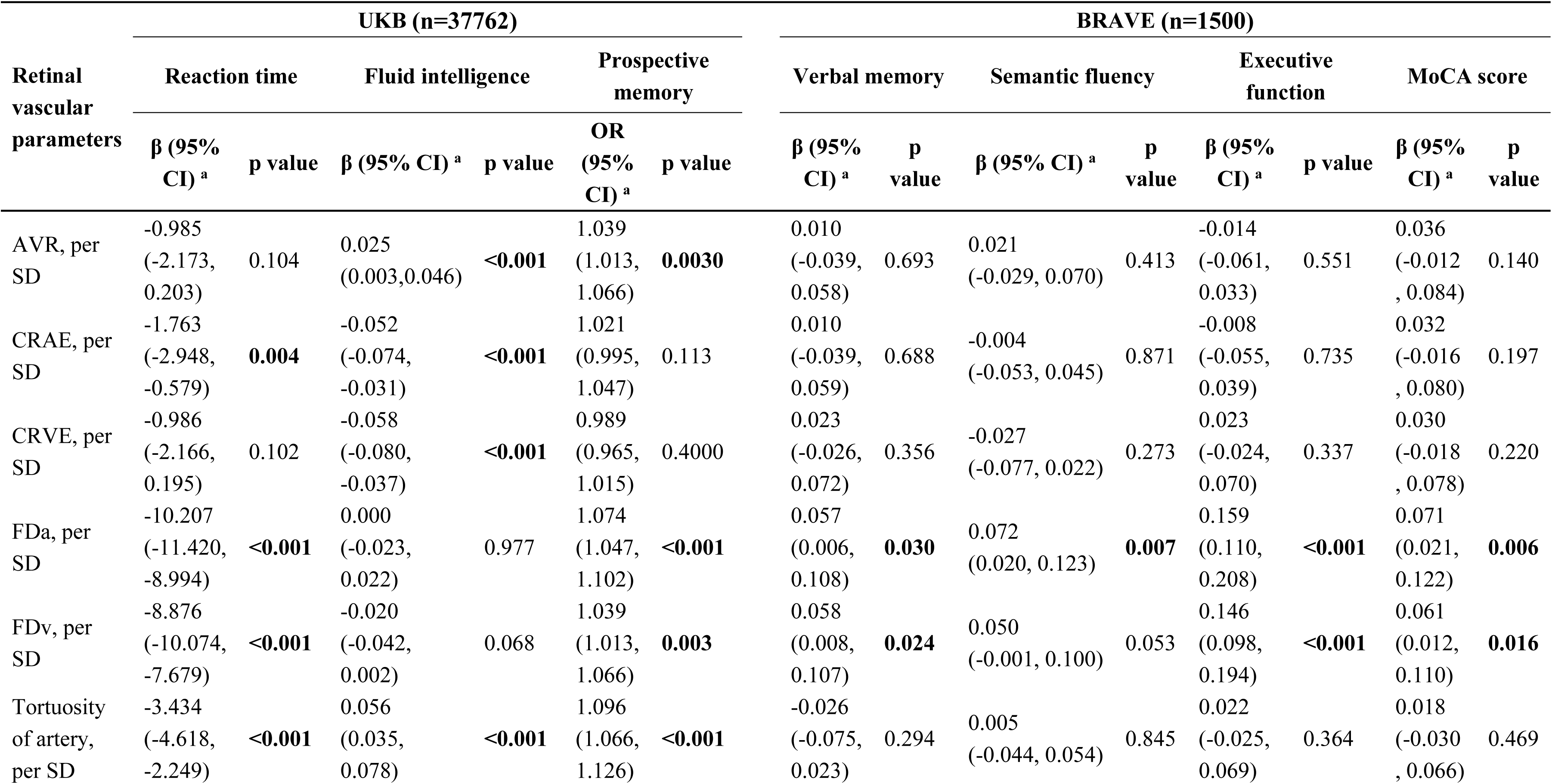
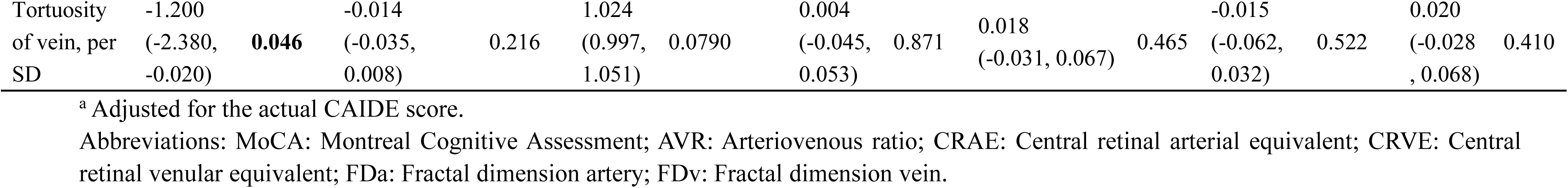
Association of retinal vascular parameters with arterial stiffness and atherosclerosis in the UKB and BRAVE study.

### Association of retinal vascular parameters with brain structure measures and dementia incidence in the UKB study

Among the four brain structure measures, the higher CRAE, FDa and FDv were consistently associated with larger volumes of the peripheral cortical gray matter, gray matter, and total brain (all p<0.05). Additionally, elevated tortuosity of vein was associated with increases in the three brain structure measures, excluding gray matter. The detailed results are summarized in the **appendix (pp 31-32)**.

We analyzed the associations between retinal vascular parameters and dementia incidence (**appendix pp 33-36**). Higher FDa and FDv were inversely associated with all-cause dementia and Alzheimer’s disease. Additionally, compared with participants in the lowest quintile of AVR, those in the highest quintile showed a lower risk of vascular dementia incidence (HR=0.294, 95% CI: 0.121-0.710).

### Model performance with the addition of retinal vascular parameters to the algorithm-estimated CAIDE model via fundus images

In the above analysis, we identified associations among the retinal vascular parameters and various domains, including arteriosclerosis, cognitive performance, brain structure measures, and dementia incidence. Furthermore, considering that our fully automated quantitative analysis algorithm allows for the acquisition of these seven retinal vascular parameters without incurring additional costs, we incorporated these parameters as quintiles into the algorithm-estimated CAIDE model to evaluate the model’s performance.

**Table 4** presents the predictive performance metrics of the algorithm-estimated CAIDE model for 14-year all-cause dementia and Alzheimer’s disease risk prediction, extended by the retinal vascular parameters. In the derivation set (n=38,384), the integration of seven retinal vascular parameters significantly improved the AUC of the algorithm-estimated CAIDE model from 0.692 (0.674-0.711) to 0.711 (0.693-0.729) for all-cause dementia (DeLong’s test p<0.001) and from 0.705 (0.677-0.732) to 0.727 (0.700-0.753) for AD (DeLong’s test p=0.014). In the internal validation set (n=9594), these findings were confirmed, showing similar improvements in the AUC, with the AUC increasing from 0.682 (0.642-0.721) to 0.706 (0.666-0.746) for all-cause dementia (DeLong’s test p=0.018) and from 0.675 (0.620-0.729) to 0.705 (0.650-0.759) for AD (DeLong’s test p=0.048).

**Table 4.**
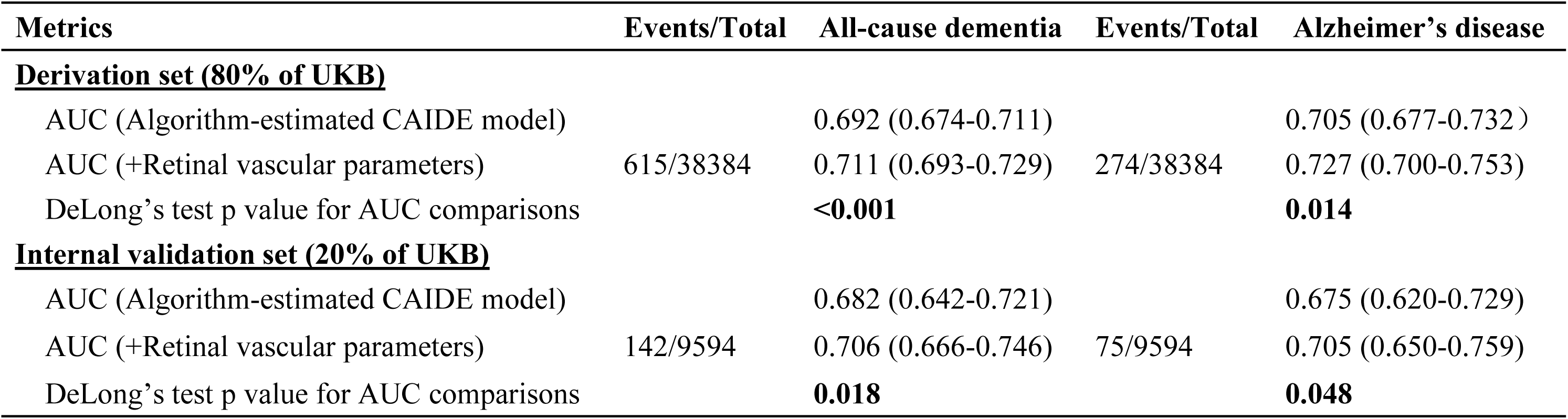
Metrics of the predictive performance of the algorithm-estimated CAIDE model for 14-year dementia risk via fundus images without and with extension by retinal vascular parameters.

| **Metrics** | **Events/Total** | **All-cause dementia** | **Events/Total** | **Alzheimer's disease** |
| --- | --- | --- | --- | --- |
| **<u>Derivation set (80% of UKB)</u>** |  |  |  |  |
| AUC (Algorithm-estimated CAIDE model) |  | 0.692 (0.674-0.711) |  | 0.705 (0.677-0.732) |
| AUC (+Retinal vascular parameters) | 615/38384 | 0.711 (0.693-0.729) | 274/38384 | 0.727 (0.700-0.753) |
| DeLong's test p value for AUC comparisons |  | **<0.001** |  | **0.014** |
| **<u>Internal validation set (20% of UKB)</u>** |  |  |  |  |
| AUC (Algorithm-estimated CAIDE model) |  | 0.682 (0.642-0.721) |  | 0.675 (0.620-0.729) |
| AUC (+Retinal vascular parameters) | 142/9594 | 0.706 (0.666-0.746) | 75/9594 | 0.705 (0.650-0.759) |
| DeLong's test p value for AUC comparisons |  | **0.018** |  | **0.048** |

We further compared the predictive performance of the algorithm-estimated CAIDE model that incorporates retinal vascular parameters with that of the original CAIDE model about the 14-year risk of dementia and its subtypes (**appendix pp 37-38**). The extended model based on fundus images displayed a pronounced improvement in the discrimination of all-cause dementia and AD, reaching statistical significance in the overall sample and males (all DeLong’s test p<0.01), but not in females (p=0.059). However, in females, the algorithm-estimated CAIDE model integrating retinal vascular parameters significantly improved discriminative ability for predicting AD and VD (p=0.014 and 0.012, respectively).

## Discussion

Improving the accessibility and ease of routine screening of high-risk groups of dementia addresses a major public health need in healthcare management. However, the commonly available risk assessment tools that require cognitive tests or multidimensional risk factors pose challenges for their application in population-based settings. In this study, we developed a deep learning algorithm for the fully automated quantification of retinal vascular parameters and analyzed over 50,000 retinal fundus images across the Chinese and British community populations. We demonstrated significant associations of retinal vascular parameters with markers of arteriosclerosis, cognitive performance, and brain MRI of neurodegeneration. The most important finding of our study is that the addition of retinal vascular parameters to the algorithm-estimated CAIDE model via fundus images enhanced the predictive ability for 14-year dementia risk, and a direct comparison with the actual CAIDE model also suggested superior predictive performance of dementia. Taken together, our study revealed the potential of using automatically extracted retinal fundus image-derived vascular parameters as biomarkers and clarified the feasibility of adopting the algorithm-estimated CAIDE model, which integrated retinal vascular parameters to screen individuals at high risk for dementia in primary healthcare settings.

To the best of our knowledge, this study is the first work to validate and enhance the predictive ability of the algorithm-estimated CAIDE model based on fundus photographs in a longitudinal cohort study with a large sample size. Emerging studies have endeavored to develop AI deep learning algorithms to detect or predict vascular-related disease utilizing retinal fundus images.^38–40^ Vascular diseases, particularly microvascular damage in the brain, are recognized as significant contributors to dementia.^41,42^ However, in the field of neurodegenerative diseases, most existing learning algorithms based on fundus images predominantly concentrate on constructing diagnostic classification models using cross-sectional data.^38^ Research investigating the potential of retinal image analysis to predict the risk of developing dementia remains scarce. In our previous study,^9^ our research group pioneered the development of a deep learning algorithm trained on over 250,000 fundus photographs to estimate the CAIDE dementia risk score. The CAIDE scores estimated by the algorithm demonstrated a high consistency with the scores obtained through traditional CAIDE model calculations, achieving a high AUC of 0.926 in an external validation for identifying individuals with high dementia risk. However, due to the lack of a longitudinal dataset that includes baseline fundus images and subsequent dementia events, the algorithm has not been validated for predicting actual dementia occurrences. In this study, we address this research gap using data from the UKB study, concluding that the performance of the algorithm-estimated CAIDE score is comparable to that of the traditional CAIDE model in predicting dementia.

The CAIDE dementia risk score is one of the most widely utilized risk scores to predict dementia; however, it exhibited only moderate predictive accuracy in this study. Similarly, in three previous studies that validated the CAIDE model in the UKB dataset, AUCs or C statistics did not reach 0.7 either.^20,21,43^ It might be due to the shorter follow-up period and relatively lower incidence rate of dementia in the UKB study.^43^ Furthermore, the CAIDE risk score was developed nearly two decades ago and therefore does not include the latest insights regarding risk factors. In recent years, several studies have attempted to extend the CAIDE model by incorporating polygenic risk scores (PRS),^5^ measures of insulin resistance,^44^ inflammation-related biomarkers,^45^ cognitive test scores,^5^ and mid-life risk factors^6^. The addition of some of these indicators had the potential to enhance the performance of the CAIDE model. For instance, in the study conducted by Trares and colleagues, the extension of the CAIDE model to include PRS resulted in an improvement of the AUC for predicting all-cause dementia, increasing from 0.785 to 0.790.^5^ Although these results are encouraging, the high time and economic costs, along with the limited accessibility of biomarkers, currently restrict the applicability of these enhanced models in primary healthcare settings. In contrast, the algorithm-estimated CAIDE model and retinal vascular parameters utilized in this study were extracted from the same fundus photograph of an individual, which means that the additional biomarkers would not increase the complexity of data collection in clinical practice. Moreover, non-mydriatic fundus photography of both eyes is a non-invasive, low-cost, and rapid examination that can be performed by a nonprofessional without the need for laboratory test resources.^39,46^ As this technology gradually becomes more widely adopted in rural areas and primary healthcare settings, our enhanced algorithm-estimated CAIDE model provides a viable technological solution for opportunistic screening of individuals at high risk for dementia, alongside the implementation of ophthalmic disease screening.

The results of the association analysis further validated the rationale for incorporating retinal vascular parameters to improve the predictive power of the CAIDE model. This study systematically investigated the associations between retinal vascular parameters and various indicators of arteriosclerosis, cognitive function, and brain MRI biomarkers across different cohorts in two countries. Notably, we found that greater FD may be indicative of a healthier vascular state, better cognitive function, and less neurodegeneration, which is consistent with several existing studies.^47,48^ However, discrepancies among various studies regarding the associations between other retinal vascular parameters and health outcomes suggest that the relationship between retinal vascular parameters and cardiovascular as well as neurodegenerative diseases is complex and may be influenced by demographic factors such as age, sex, race, and comorbidities.^47,49^ Further large-scale and longitudinal studies are needed to clarify these associations more thoroughly. We also noted that many previous studies have been conducted with small sample sizes and relied on semi-automated analysis of retinal vascular parameters.^50,51^ Semi-automated computer software has achieved measurement of various retinal vascular characteristics.^47,52^ Nevertheless, semi-automated approaches, which require manual intervention, are inefficient in large sample analysis. The fully automated algorithm, which removes the need for manual intervention, makes it feasible to obtain quantitative measures of vessel morphology in large-scale epidemiological studies.^10,53^ Utilizing the AI algorithm, fundus photographs can be swiftly processed for objective metric extraction, contrasting the subjective, error-prone, labor-intensive, and consequently time-consuming manual method.^53^ However, performance of A/V classification of the proposed algorithm is slightly inferior to the automatic analysis method developed by Ma et al., which is so far the optimal method with a A/V classification accuracy of 94.50%.^24^ The high accuracy may be attributed to the adopted deep network with a spatial activation mechanism that performs full vessel segmentation and A/V classification simultaneously, so the accuracy of both tasks could be improved.^24^ However, this method is complex and time-consuming. Apart from that, the algorithm developed by our research group demonstrates better robustness and is capable of adapting to a greater variety of real-world low-quality and lesion-rich fundus images. With all the advantages mentioned above, our fully automated analysis methods could enable us to quantify retinal vascular characteristics in a large sample population in communities or even nationwide.

The major strengths of our study included the large sample size of international community-based participants, the application of a fully automated algorithm to measure retinal vascular parameters, the assessment of multiple markers of arteriosclerosis, cognitive function, and brain MRI measures, as well as valuable follow-up records of dementia events. However, several potential limitations should be noted. Firstly, we did not test the robustness of the algorithm-estimated CAIDE model integrating retinal vascular parameters in the external validation cohorts due to the lack of other available perspective cohorts with baseline fundus photographs and follow-up dementia events. Secondly, the original CAIDE score was designed for 20-year dementia risk prediction. However, we only focused on the 14-year dementia risk due to the lack of sufficient follow-up in the UKB study. Thirdly, when comparing the original CAIDE model with the algorithm-estimated CAIDE model, we excluded individuals with missing variables in the original CAIDE model to better align with clinical practice. In exploring whether the addition of retinal vascular parameters to the algorithm-estimated CAIDE model could enhance its predictive ability, the sample size for analysis increased from 35,838 to 47,978 individuals, as this analysis did not involve the original CAIDE model. If there was a significant difference in dementia risk between individuals excluded due to missing variables in the original CAIDE model and those included in the analysis, this could affect the actual performance of the model. Nevertheless, it also reflects the challenge of obtaining complete CAIDE model variables, even in high-quality, large cohort studies like the UKB study. Therefore, the enhanced algorithm-estimated CAIDE model utilizing fundus images offers a significant advantage in large-scale research.

## Conclusion

By automatically quantifying the retinal vascular parameters using a deep learning algorithm, this study provides initial evidence of the retinal vascular biomarkers to improve dementia prediction. Compared to the original CAIDE model, the retinal vascular parameters-enhanced algorithm-estimated CAIDE model may provide a more accurate dementia risk assessment with just a single fundus photograph. Our automated approach not only streamlines screening procedures but also holds promise for implementation in primary healthcare settings, facilitating early intervention strategies. However, due to the lack of external validation, large-scale real-world evaluations of this model require further investigation.

## Data Availability

Data from the UK Biobank (http://www.ukbiobank.ac.uk/) are available to researchers upon application.

## Contributors

Wuxiang Xie, Fanfan Zheng, Darui Gao, Yanyu Zhang, and Jianhao Xiong conceptually designed the study. Darui Gao, Yanyu Zhang, Jianhao Xiong, and Sijin Zhou collected and analysed the data. Wuxiang Xie, Darui Gao and Yanyu Zhang performed the statistical analysis. Darui Gao and Yanyu Zhang drafted the manuscript. Wuxiang Xie and Fanfan Zheng had full access to all the data in the study and had responsibility for the integrity of the data and the accuracy of the data analysis. All authors reviewed the manuscript and were responsible for the decision to submit the manuscript. Darui Gao, Yanyu Zhang, and Jianhao Xiong contributed equally and shared co-first authorship. Wuxiang Xie and Fanfan Zheng jointly supervised this work and shared the co-last authorship.

## Data sharing

Data from the UK Biobank (http://www.ukbiobank.ac.uk/) are available to researchers upon application. This research was conducted using the UK Biobank Resource under Application 90492.

## Acknowledgments

We gratefully appreciate all of the contributions of the research participants, clinicians and imaging and laboratory technicians.

