## Supplementary appendix for "A deep learning algorithm based on fundus photographs to measure retinal vascular parameters and their additional value beyond the CAIDE risk score for predicting 14-year dementia risk"

**Catalogue**

Supplementary Method 2: The detailed methods for the development and evaluation of the algorithm for the segmentation and quantification of retinal vascular parameters16

**Supplementary Method 1: Study design, datasets, and participants**

We conducted a multistage population-based study in five parts as described later, using prospective and cross-sectional datasets. The overall study design is described in **Figure 1**.

Firstly, we used a published deep learning algorithm[1] developed by our research group for estimating the CAIDE dementia risk score via fundus images to obtain an algorithm-estimated score for each participant with qualified fundus photographs in the UK Biobank (UKB) study (Application Number: 90492). The UKB study recruited over 0.5 million participants aged 40 to 60 years from the United Kingdom between 2006 and 2010, with detailed information reported previously.[2] Ethics Committee approval for the UK Biobank was obtained from the North West Multi-Centre Research Ethics Committee (Research Ethics Committee reference: 16/NW/0274). Informed consent was obtained from all participants. The UKB eye and vision substudy, which commenced in late 2009 at six assessment centers as an enhancement to the initial baseline measures, included retinal imaging scans conducted during participants' visits.[3] Our analysis was based on 97,949 retinal fundus images (Field ID: 21016 and 21015) of 61,418 subjects captured during the initial assessment visit (Instance 0). To compare the algorithm-estimated CAIDE score with the traditional CAIDE model computed score, a combination of variables derived from the participants' questionnaire and blood samples was also collected.

Secondly, to develop the automatic quantitative analysis algorithm for vascular network parameters measurement, the training dataset included 591 retinal color fundus images with labeling of vessel segmentation and artery/vein (A/V) classification provided. Of these, 40 images were from the publicly available AV-DRIVE database.[4] The remaining images were sourced from our internal Chinese medical check-up database, and each image was manually labelled by 2 experienced technicians. To evaluate the proposed algorithm, 269 fundus images from our internal database and AV-DRIVE database were used.

Thirdly, the algorithm mentioned above was applied to the fundus images collected by the UKB study and the Beijing Research on Ageing and Vessel (BRAVE) study for the segmentation and quantification of retinal microvasculature. The BRAVE study is a community-based, prospective cohort study investigating the contributions of vascular conditions to cognitive impairment and dementia. In 2019, all 1789 residents aged 40–80 years from the Xishan community, Shijingshan District, were invited to participate in the baseline survey.[5] The study was approved by the Institutional Review Board of Peking University Health Science Center (IRB0001052-19060) and Fuwai Hospital (IRB2012-BG-006), and all participants gave their written informed consent according to the Declaration of Helsinki. A total of 1554 participants were initially enrolled and underwent baseline evaluation. Of these, 3191 fundus images were collected from 1530 subjects during baseline assessments. In the BRAVE study, all fundus images were acquired using the same Centervue DRS automatic fundus camera. All images were captured using 45° fields of view.

Fourthly, we investigated the associations between retinal vascular parameters with arterial stiffness, atherosclerosis, and cognitive performance in the UKB and BRAVE studies, respectively. Furthermore, in the UKB study, we conducted a further analysis of the relationship between retinal vascular parameters and brain structural measures and dementia incidence.

Finally, the UKB dataset was randomly split into a derivation (80%) and a validation set (20%) to assess whether 14-year dementia risk prediction with the estimated CAIDE dementia risk score via fundus images can be improved by incorporating retinal vascular parameters. Additionally, we further assessed whether the improved estimated CAIDE dementia risk score derived from fundus images demonstrates superior predictive performance for 14-year dementia risk compared to the traditional CAIDE model computed risk score in the UKB dataset.

**Supplementary Figure 1: Flowchart of participant selection for stage 1**

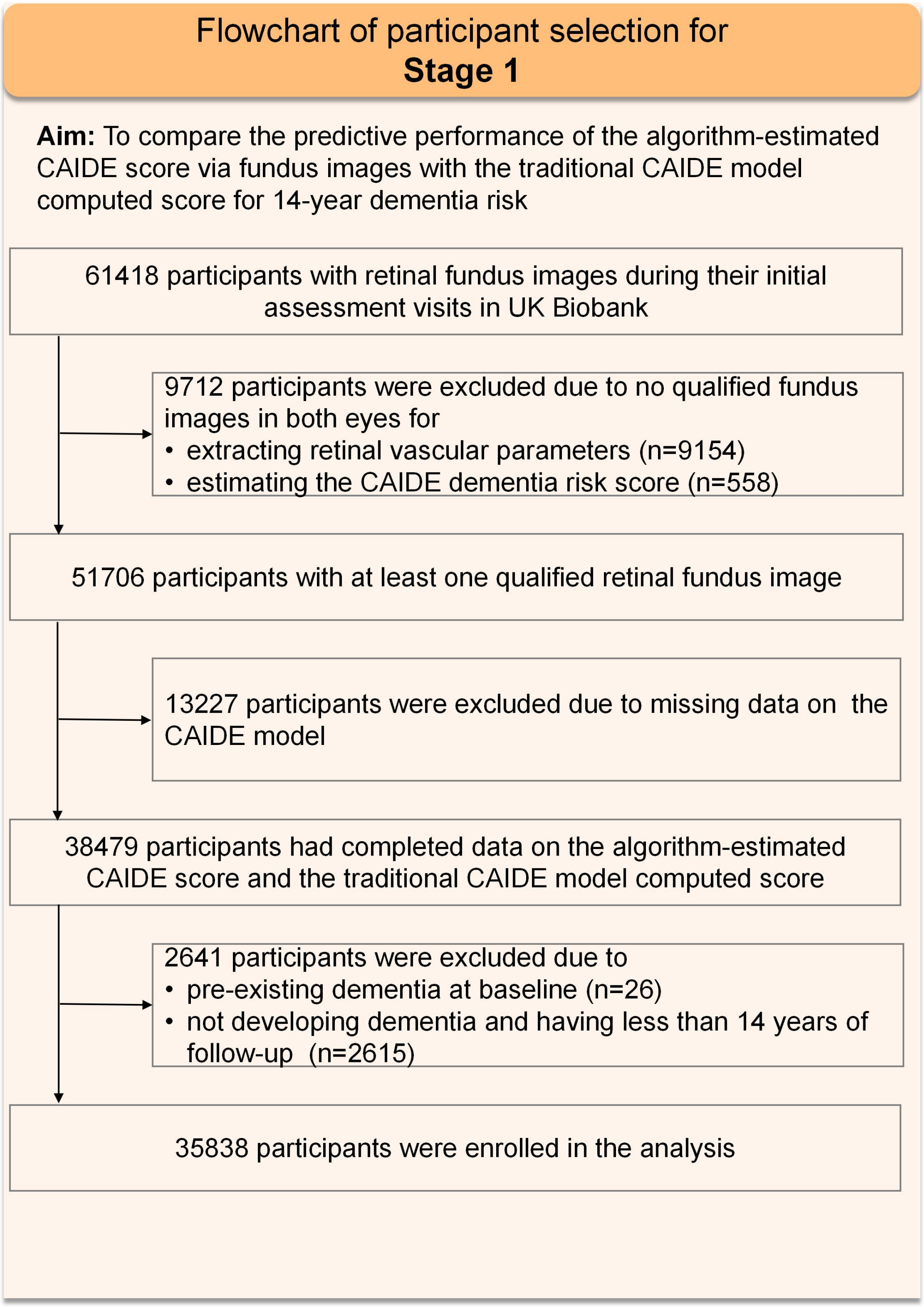

**Supplementary Figure 2: Flowchart of participant selection for stages 2-4 in the UKB dataset**

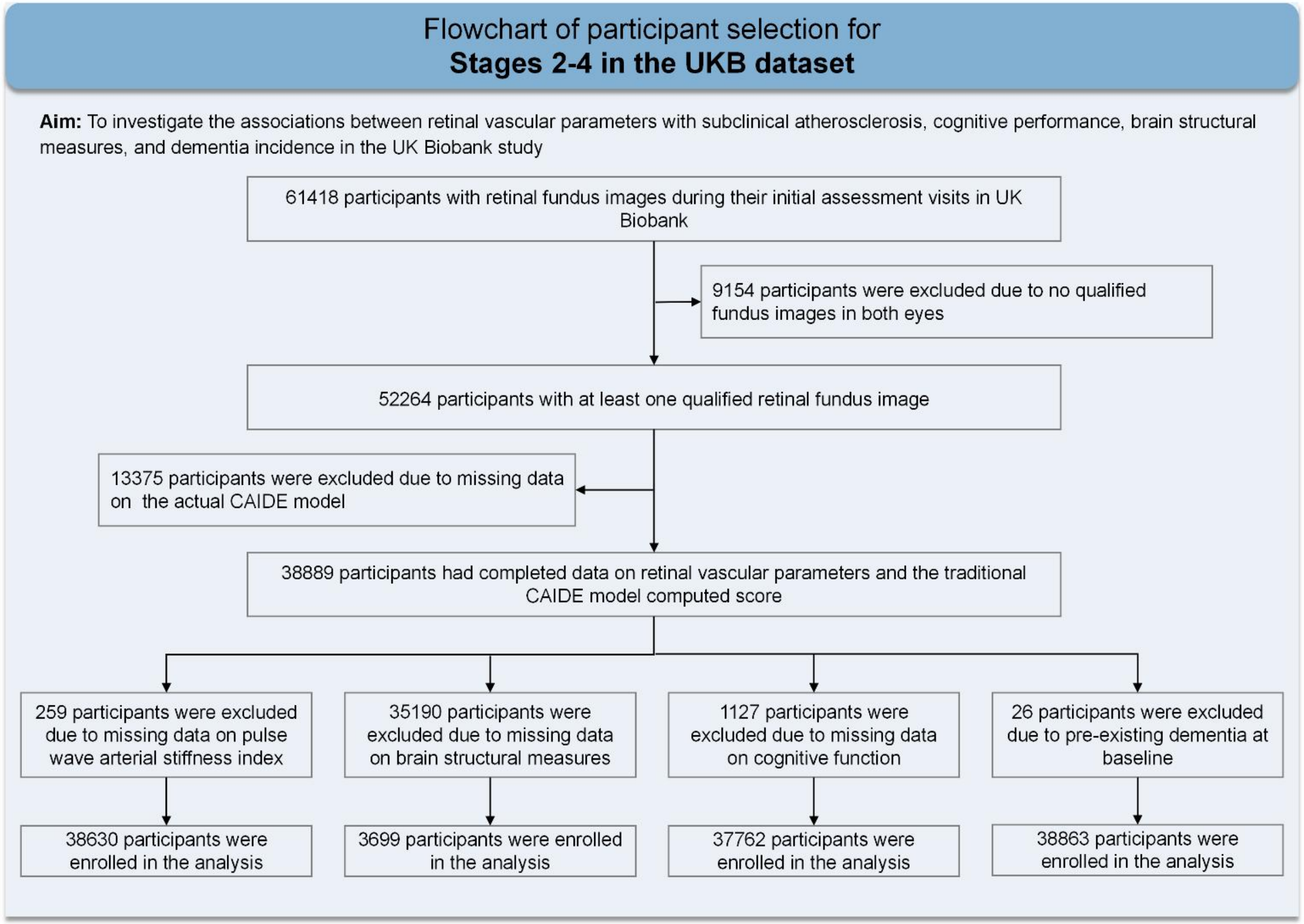

**Supplementary Figure 3: Flowchart of participant selection for stages 2-4 in the BRAVE dataset**

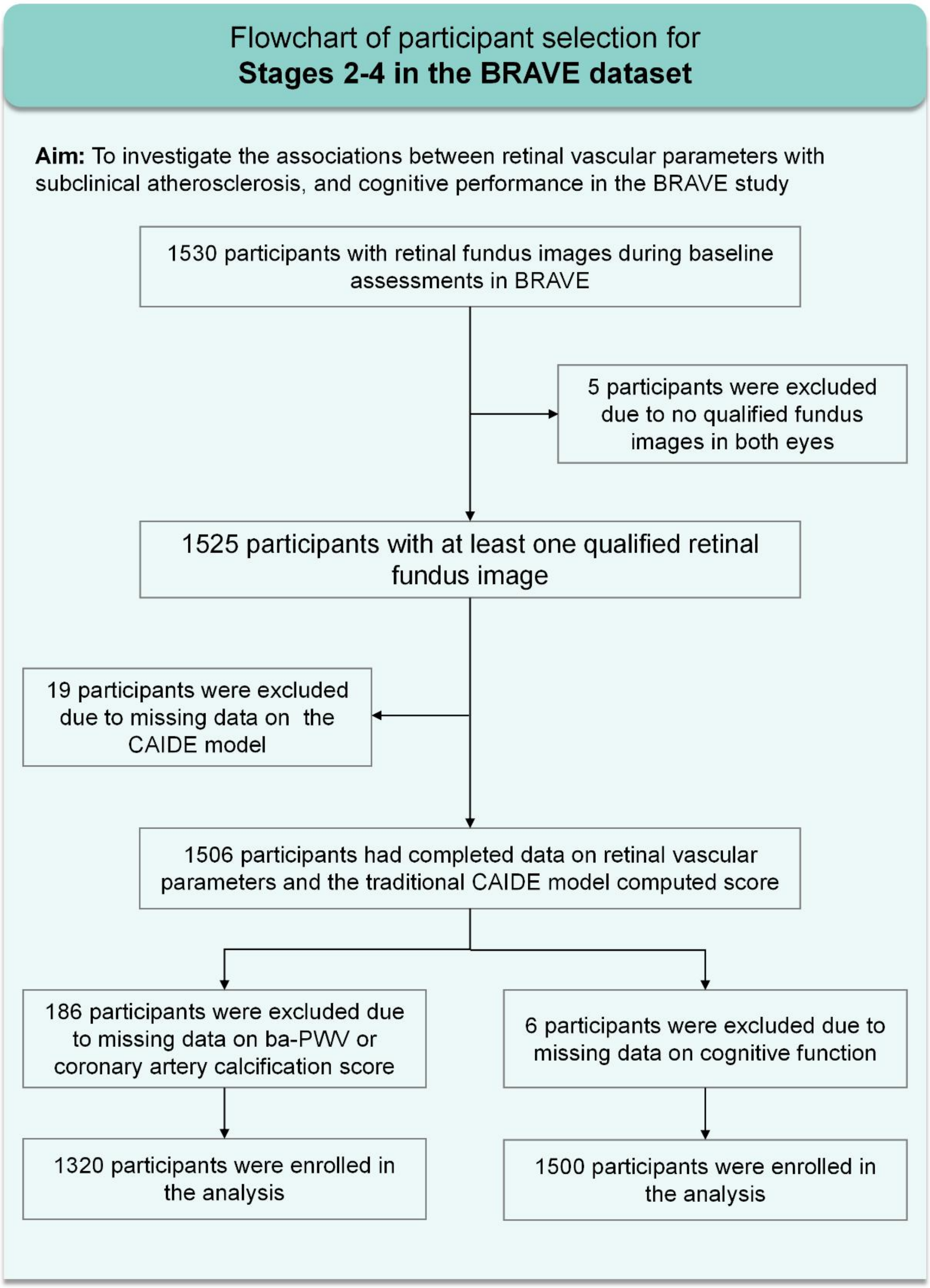

### Supplementary Figure 4: Flowchart of participant selection for stage 5

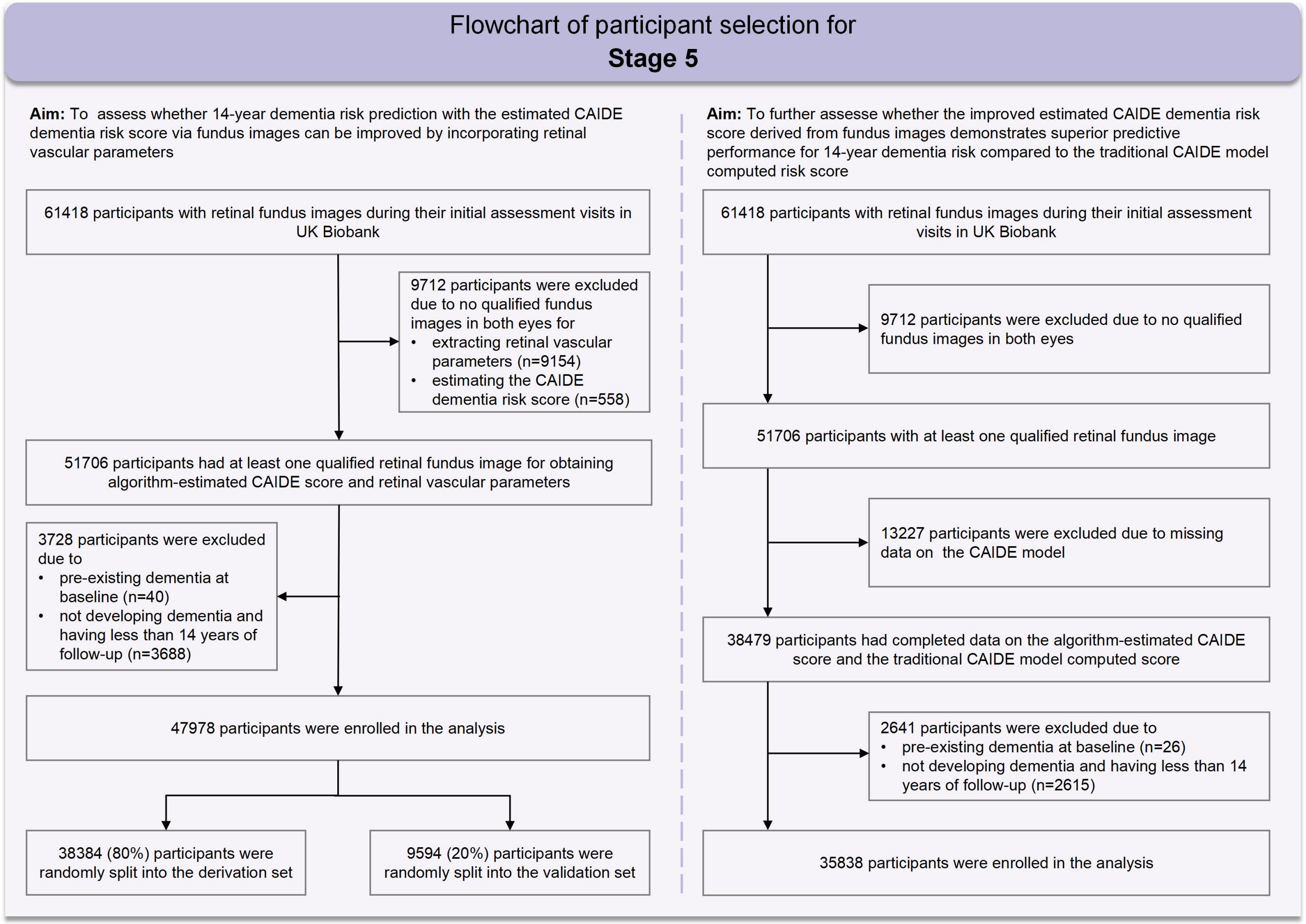

**Supplementary Table 1: Baseline characteristics of selected participants from the UKB study and the BRAVE study Supplementary**

| **Baseline characteristics** | **UK Biobank (N=35838)** | **BRAVE (N=1506)** |
|---|---|---|
| Age at enrollment, years | 56.4 ± 8.2 | 59.8 ± 7.2 |
| Male | 16445 (45.9) | 561 (37.3) |
| Years of education | | |
| ≥10 | 25834 (72.1) | 901 (59.8) |
| 7-9 | 5921 (16.5) | 512 (34.0) |
| 0-6 | 4083 (11.4) | 93 (6.2) |
| Systolic blood pressure, mm Hg | 136.2 ± 18.2 | 132.5 ± 17.1 |
| Body-mass index, kg/m2 | 27.1 ± 4.6 | 26.0 ± 3.4 |
| Total cholesterol, mmol/L | 5.7 ± 1.1 | 4.9 ± 1.0 |
| Inactive physical activity | 6217 (17.3) | 1235 (82.0) |

**Supplementary Table 2: Definition of the CAIDE model variables**

| | | UK Biobank | | BRAVE |
|---|---|---|---|---|
| Variables | Category | Description | Data-Field ID | Description |
| Age | <47 = 0 points,<br>47-53 = 3 points<br>>53 = 4 points | Age in years: Difference between date attended baseline assessment and date of birth recorded by NHS | 53, 34 | Age in years: According to the self-reported questionnaire or the registered ID card information to calculate. |
| Sex | female = 0 points<br>male = 1 point | Sex of participant: NHS derived and/or touchscreen questionnaire. | 31 | Based on information derived from self-reported questionnaires. |

| | | | | |
|---|---|---|---|---|
| Systolic blood pressure | ≤140mmHg = 0 point<br>>140mmHg = 2 points | Two automated readings (a few moments apart) of systolic blood pressure were taken. The mean across these two measurements were used, unless one measurement was missing. | 4080 | For each participant, brachial blood pressure was measured 3 times in a seated position after at least 5minutes of rest, and the average of the second and third readings was used in the analysis. |
| Body-mass index | ≤30 = 0 points<br>>30 = 2 points | Calculated from height and weight collected at baseline. | 21001 | Calculated from height and weight collected at baseline. |
| Total cholesterol | <6.5mmol/l = 0 points<br>>6.5mmol/l = 1 points | Blood samples (non-fasting) were collected and assayed with a Beckman Coulter AU5800 analytical platform to determine total, HDL and LDL levels. | 30690 | Blood samples (fasting) were collected. |

| | | | | |
|---|---|---|---|---|
| Physical activity | active = 0 points<br>inactive = 1 point | IPAQ classifications of physical activity groups were used, with "moderate" to "high" activity groups defined as "active", and "low" defined as "inactive". | 22032 | According to the self-reported questionnaire: "Active": Engaging in vigorous physical activity at least twice a week (such as sports that induce an increased heart rate and sweating, like badminton, tennis, and soccer), with each session lasting a minimum of 20 minutes. Conversely, individuals not meeting this criterion are classified as "inactive". |

| | | | | |
|---|---|---|---|---|
| Education | >10 = 0 points<br>7–9 = 2 points<br>0–6 = 3 points | Years of education: Which of the following qualifications do you have: college or university degree; A level/AS levels or equivalent; O levels/General Certificate of Secondary Education (GCSE) or equivalent/Certificate of Secondary Education (CSE) or equivalent; National Vocational Qualification (NVQ), Higher National Diploma (HNC), or equivalent; other professional qualifications (e.g., nursing, teaching); none of the above.<br>We map each educational qualification to an International Standard Classification of Education (ISCED) category and impute years of education equivalent for each ISCED category. The imputed years of education for the educational qualifications are: no qualification = 6 years; CES/O levels/GCSEs or equivalent = 9 years; A level/AS levels or equivalent = 12 years; other professional qualification = 15 years; NVQ/HNC or equivalent = 19 years; and college or university degree =20 years. | 6138 | Years of education: Based on information derived from self-reported questionnaires, years of education is defined as follows: primary education or below, junior high school, and senior high school or above, corresponding to educational attainment of 0-6 years, 7-9 years, and 10 years or more, respectively. |

**Supplementary Method 2: The detailed methods for the development and evaluation of the algorithm for the segmentation and quantification of retinal vascular parameters**

In this paper, we measured the following vascular parameters, including central retinal arterial equivalent (CRAE), central retinal venular equivalent (CRVE), arteriovenous ratio (AVR), fractal dimension artery (FDa), fractal dimension vein (FDv), tortuosity of artery and tortuosity of vein. Next, we will describe the calculation methods of each parameter in detail.

Firstly, we use the coarse segmentation model of arteries and veins. This model utilizes an U-Net based deep learning segmentation network to automatically extract the vascular structures from retinal fundus images upon input, yielding a mask containing foreground information of arteries and veins. Then we use an object detection model which uses the network structure of Yolov3. The model takes retinal fundus images as input for target detection of optic disc and macula, resulting in positional information of the optic disc and macula. Based on this positional information, the retinal vessel measurement and analysis system automatically identifies four regions within the retina: temporal superior, temporal inferior, nasal superior, and nasal inferior. The foreground masks of arteries and veins, outputted by the coarse segmentation model, were normalized to a size of 512×512. Subsequently, the box-counting dimension algorithm was applied with a box size ranging from 10 to 200 and a step size of 5 to compute the fractal dimensions of arteries and veins. Secondly, we delineate the regions of retinal vessel caliber measurement on images that have undergone arteriovenous segmentation as well as macula and optic disc detection. Specifically, it can be divided into the following steps.

(1) Based on the detected bounding box information of the optic disc, the diameter value D of the optic disc is calculated. Subsequently, an annular region with a radius ranging from 1 times to 1.5 times the diameter of the optic disc is defined as the optic disc B-zone (RB);

(2) The coarse segmentation masks of both arteries and veins were individually

processed to obtain their respective skeletons. After setting the optic disc region to zero in the skeleton mask, each independent connected component was extracted, and its centroid was calculated. The connected regions of arteries and veins, where the centroids were distributed within the temporal superior/inferior regions, were separately evaluated. From these regions, the main vascular branch was identified by selecting the connected component with the largest foreground area.

(3) Within the RB region, the central points P of the main vascular branch skeletons of the arteries and veins in the temporal superior/inferior regions were obtained by calculating the vascular segments. Subsequently, regions of interest (ROI) were defined by selecting an area centered at point P with a length and width equal to 2 times the diameter of the optic disc D.

Subsequently, a fine segmentation model for arteries and veins was utilized to perform individual segmentation on image patches of the four ROI. This segmentation process yielded high-precision foreground masks for arteries and veins within each respective image patch. The fine segmentation model is based on an U-Net architecture deep learning segmentation network, which was trained with supervised learning using image patches of the regions of interest annotated with arterial and venous segmentation labels. The input size is a 640×640 RGB image, and the output size is a 640×640 mask that contains high-precision foreground information of both arteries and veins.

After obtaining the high-precision foreground information of arteries and veins, the centroid point P of each region of interest and the corresponding localized arterial and venous segmentation masks were extracted to obtain the target vascular segments for measurement. Subsequently, existing skeleton extraction algorithms were applied separately to the segmentation masks of arteries and veins to obtain skeleton masks representing the central regions of the blood vessels. Then, established distance calculation algorithms were employed to obtain the diameter values of each central point of the blood vessels. The average diameter value of each central point in the target vascular segment was calculated as the diameter value of that particular segment. Lastly, the diameter ratio of arteries and veins was computed.

The measurement of tortuosity involves several steps. Firstly, the connected regions of arteries and veins that exceed a predefined threshold are subjected to skeletonization. Subsequently, starting from the intersection point at the edge of the optic disc, a depth-first search algorithm is employed to traverse the blood vessels and separate them into the first, second, third, and fourth branching orders. Then, for each individual vessel, the curvature is estimated using a curvature estimation method. The average tortuosity value of the arteries and veins is computed by taking the mean of the individual curvature values.

We evaluated our proposed algorithm with two different tasks. The first task was conducted on our internal image dataset. Initially, we randomly selected 249 fundus images from our internal database, which were independently labelled by three experienced technicians. The labelled results were recorded as the standard test dataset. We used mean Intersection over Union (mIoU) as the metric to evaluate the performance of segmentation. The mIoU, which is a widely adopted metric in image segmentation evaluation, quantifies the overall concordance between the segmented output of an algorithm and the truth labels.[6,7] The mIoU is optimal at a value of 1 and worst at a value of 0. We first calculated the IoU between three technicians, took the average, and obtained the human-human mIoU. Then, the average of the IoU between the algorithm and the three technicians was calculated, obtaining the human-algorithm mIoU. Finally, we compared the human-human mIoU and the human-algorithm mIoU to verify whether the proposed algorithm reached the human average level. In addition, we evaluated the proposed algorithm on the AV-DRIVE database to verify the robustness. In this external evaluation, we adopted four metrics for the evaluation of vessel segmentation: the average accuracy (Acc), sensitivity (Sen), specificity (Spe), and area under curve (AUC).[8] A/V classification performance was evaluated using pixel-wise Acc, Sen and Spe for the ground-truth artery vein pixels.[8] By taking arteries as positives and veins as negatives, Sen reflects the proposed algorithm's capability of detecting arteries and Spe for veins.

By utilizing this algorithm, we achieved fully automated quantitative measurement of seven meaningful parameters of the retinal vascular network.

Specifically, the extracted parameters included CRAE, CRVE, AVR, FDa, FDv, tortuosity of artery, and tortuosity of vein.

**Supplementary Table 3: Descriptions of retinal vascular parameters**

| Retinal vascular network parameters | Description | Reference |
|---|---|---|
| Arteriovenous ratio (AVR) | The ratio of arterial to venous diameter:<br>AVR=CRAE/CRVE | Frost et al.[9] |
| Central retinal arterial equivalent (CRAE)<br>Central retinal venular equivalent (CRVE) | Conducted an overall measurement of the equivalent diameter of individual blood vessels or the six largest blood vessels based on the Knudston-Parr-Hubbard formula.[10] | Williams et al.;[11]<br>Wong et al.;[12]<br>de Jong et al. ;[13]<br>Frost et al.;[9]<br>Cheung et al.;[14]<br>Seidelmann et al..[15] |
| Tortuosity of artery<br>Tortuosity of vein | The path of blood vessel trajectory was integrated and the length of the vessels was standardized, reflecting the tortuosity of the blood vessels. | Williams et al.;[11]<br>Frost et al.;[9]<br>Cheung et al..[14] |
| Fractal dimension artery (FDa)<br>Fractal dimension vein (FDv) | A measurement was performed to assess the overall complexity of the retinal vascular network, reflecting its two-dimensional fractal pattern. A higher numerical value indicates greater complexity of the fractal structure. The box-counting method was employed for the calculation. | Williams et al.;[11]<br>Frost et al.;[9]<br>Cheung et al..[14] |

**Supplementary Table 4: ICD-9 & ICD-10 codes used to ascertain all-cause and cause-specific dementia cases**

| UK Biobank Self Report Codes | | | | | | |
|---|---|---|---|---|---|---|
| **Code Type** | **Code** | **Biobank Code Text** | **AD** | **VD** | **FTD** | **Dementia** |
| UK Biobank Self Report | Field 20002 Code 1263 | Dementia/Alzheimer's/Cognitive Impairment | | | | √ |
| **ICD 9 Codes** | | | | | | |
| **Code Type** | **ICD 9 Code** | **ICD 9 Text** | **AD** | **VD** | **FTD** | **Dementia** |
| ICD 9 Code | 290.2 | Senile dementia, depressed or paranoid type | | | | √ |
| ICD 9 Code | 290.3 | Senile dementia with acute confusional state | | | | √ |
| ICD 9 Code | 290.4 | Arteriosclerotic dementia | | √ | | √ |
| ICD 9 Code | 291.2 | Other alcoholic dementia | | | | √ |
| ICD 9 Code | 294.1 | Dementia in other conditions classified elsewhere | | | | √ |
| ICD 9 Code | 331.0 | Alzheimer's disease | √ | | | √ |
| ICD 9 Code | 331.1 | Pick's disease | | | √ | √ |
| ICD 9 Code | 331.2 | Senile degeneration of brain | | | | √ |

| | | | | | | |
|---|---|---|---|---|---|---|
| ICD 9 Code | 331.5 | Creutzfeldt-Jakob disease | | | | √ |
| **ICD 10 Codes** | | | | | | |
| **Code Type** | **ICD 10 Code** | **ICD 10 Text** | **AD** | **VD** | **FTD** | **Dementia** |
| ICD 10 Code | A81.0 | Sporadic Creutzfeldt-Jakob disease | | | | √ |
| ICD 10 Code | F00 | Dementia in Alzheimer's disease | √ | | | √ |
| ICD 10 Code | F00.0 | Dementia in Alzheimer's disease with early onset | √ | | | √ |
| ICD 10 Code | F00.1 | Dementia in Alzheimer's disease with late onset | √ | | | √ |
| ICD 10 Code | F00.2 | Dementia in Alzheimer's disease, atypical or mixed type | √ | | | √ |
| ICD 10 Code | F00.9 | Dementia in Alzheimer's disease, unspecified | √ | | | √ |
| ICD 10 Code | F01 | Vascular dementia | | √ | | √ |
| ICD 10 Code | F01.0 | Vascular dementia of acute onset | | √ | | √ |
| ICD 10 Code | F01.1 | Multi-infarct dementia | | √ | | √ |

| | | | | | | |
|---|---|---|---|---|---|---|
| ICD 10 Code | F01.2 | Subcortical vascular dementia | | √ | | √ |
| ICD 10 Code | F01.3 | Mixed cortical and sub-cortical vascular dementia | | √ | | √ |
| ICD 10 Code | F01.8 | Other vascular dementia | | √ | | √ |
| ICD 10 Code | F01.9 | Vascular dementia, unspecified | | √ | | √ |
| ICD 10 Code | F02 | Dementia in other diseases classified elsewhere | | | | √ |
| ICD 10 Code | F02.0 | Dementia in Picks disease | | | √ | √ |
| ICD 10 Code | F02.1 | Dementia in Creutzfeldt-Jacob disease | | | | √ |
| ICD 10 Code | F02.2 | Dementia in Huntington's disease | | | | √ |
| ICD 10 Code | F02.3 | Dementia in Parkinson's disease | | | | √ |
| ICD 10 Code | F02.4 | Dementia in HIV disease | | | | √ |
| ICD 10 Code | F02.8 | Dementia in other specified diseases is classified elsewhere | | | | √ |
| ICD 10 Code | F03 | Unspecified dementia | | | | √ |
| ICD 10 Code | F05.1 | Delirium superimposed on dementia | | | | √ |

| | | | | | | |
|---|---|---|---|---|---|---|
| ICD 10 Code | F10.6 | Mental and behavioural disorders due to use of alcohol - amnesic syndrome | | | | √ |
| ICD 10 Code | G30 | Alzheimer’s disease | √ | | | √ |
| ICD 10 Code | G30.0 | Alzheimer’s disease with early onset | √ | | | √ |
| ICD 10 Code | G30.1 | Alzheimer’s disease with late onset | √ | | | √ |
| ICD 10 Code | G30.8 | Other Alzheimer's disease | √ | | | √ |
| ICD 10 Code | G30.9 | Alzheimer's disease unspecified | √ | | | √ |
| ICD 10 Code | G31.0 | Circumscribed brain atrophy | | | √ | √ |
| ICD 10 Code | G31.1 | Senile degeneration of brain | | | | √ |
| ICD 10 Code | G31.8 | Other specified degenerative diseases of nervous system | | | | √ |
| ICD 10 Code | I67.3 | Binswanger’s disease | | √ | | |

**Supplementary Method 3: Methods of assessment of arterial stiffness and atherosclerosis in the BRAVE study**

In the BRAVE study, brachial ankle pulse wave velocity (ba-PWV) was assessed using a non-invasive arteriosclerosis measuring device (Vascular Inspecting Apparatus MB3000, M&B Electronic Instruments, China). Measurements were performed with the subject in the supine position, following a minimum of 5 minutes of rest. Occlusion cuffs were applied to the brachial arteries of both the left and right arms, as well as the ankles. Electrodes were placed on both wrists, and a heart sound sensor was positioned on the left margin of the sternum. The device recorded ECG, phonocardiogram, pulse volume waveform, heart rate, and arterial blood pressure data simultaneously, and automatically analyzed the results.[5,16] Coronary artery calcification (CAC) was measured using a multi-slice CT scan (Brilliance iCT, Philips Healthcare, Cleveland, OH, USA). CT scans were performed during breath-holding, with a tube voltage of 120 kV and a slice thickness of 2.5 mm. The scan range extended from the bifurcation of the trachea to the bottom of the heart. All images were transferred to a workstation (Advantage Workstation version 4.6, GE Healthcare, USA) for independent evaluation of coronary calcium. CAC was quantified using the Agatston score.[17] Each calcium lesion was identified as a hyperattenuating region with a CT density exceeding 130 Hounsfield units (HU). The CAC score (CACS) was automatically calculated by manually adding all detectable calcification lesions.[18]

**Supplementary Method 4: Methods of assessment of cognitive outcomes in the BRAVE study**

In the BRAVE study, the primary cognitive measurement was the Chinese version of Montreal Cognitive Assessment Basic (MoCA-B), a sensitive and validated cognitive test battery to comprehensively assess nine cognitive domains.[19] The total MoCA-B score represents global cognitive function, and a higher score indicates better cognitive function. The Montreal Cognitive Assessment Basic evaluates 9 cognitive domains: executive function, verbal memory, semantic fluency, orientation, calculation, abstraction, visual perception, naming, and attention. The assessment takes ≈15minutes and has a maximum score of 30 points. All interviewers of the BRAVE study who were involved in the cognitive tests had attended a Training & Certification Program for the MoCA.

Furthermore, a comprehensive battery of cognitive tests was administered to establish stable measures across three individual cognitive domains. Verbal memory was evaluated through the immediate and delayed recall of ten unrelated words (0–20 points). These assessments have demonstrated good construct validity and consistency, with higher scores indicating superior performance.[20] Semantic fluency was assessed using the category fluency tests for animals and fruits from the MoCA-B. Participants were instructed to name as many animals or fruits as possible within one minute. The total score was calculated by summing the unique animals and fruits identified across both tests. The category fluency test for animals has good reliability and validity.[21] Executive function was assessed using the Trail Making Test (TMT), where participants were required to draw lines connecting consecutive numbers from 1 to 25 in ascending order (TMT-A) and to alternate between connecting numbers of different colors (pink and yellow) in TMT-B. The sum of the time (seconds) taken to complete both tasks, with a maximum of three minutes per test, was calculated and subsequently inverted to generate an executive function score, where a higher score indicates better performance.[5] All cognitive scores in the BRAVE were converted to standardized Z scores by subtracting the mean scores and dividing by the standard deviation (SD) to facilitate comparisons across tests, where a higher score indicates better cognitive performance.

**Supplementary Table 5: Consistency of the performance of the algorithm for blood vessel segmentation with human labeling**

| Method | MIoU of arteries | MIoU of veins |
|---|---|---|
| Human-human | 0.4716 | 0.5241 |
| Human-algorithm | 0.4506 | 0.5240 |

Abbreviation: MIoU, mean Intersection over Union;

**Supplementary Table 6: Performance comparison of vessel segmentation on the AV-DRIVE dataset**

| Methods | Acc (%) | Spe (%) | Sen (%) | AUC (%) |
|---|---|---|---|---|
| Fu et al.[22] | 94.7 | - | 72.94 | - |
| Liskowski et al.[23] | 95.35 | 98.07 | 78.11 | 97.90 |
| Wu et al.[24] | 95.67 | **98.19** | 78.44 | 98.07 |
| Ma et al.[8] | **95.70** | 98.11 | 79.16 | 98.10 |
| **Proposed** | 94.74 | 95.02 | **91.76** | **98.38** |

Abbreviation: Acc, average accuracy; Sen, sensitivity; Spe, specificity; AUC, area under curve

**Supplementary Table 7: Performance comparison of A/V classification on the AV-DRIVE dataset**

| Methods | Acc (%) | Sen (%) | Spe (%) |
|---|---|---|---|
| Dashtbozorg et al.[25] | 87.4 | 90.0 | 84.0 |
| Estrada et al.[26] | 93.5 | 93.0 | 94.1 |
| Xu et al.[27] | 92.3 | 92.9 | 91.5 |
| Zhao et al.[28] | - | 91.9 | 91.5 |
| Ma et al.[8] | **94.5** | **93.4** | **95.5** |
| **Proposed** | 90.1 | 89.7 | 91.8 |

Abbreviation: Acc, average accuracy; Sen, sensitivity; Spe, specificity

**Supplementary Table 8: Associations between retinal vascular network parameters with brain structure measures (n=3699)**

| Retinal vascular parameters | Peripheral cortical gray matter volume ($mm^3$) | | Gray matter volume ($mm^3$) | | White matter volume ($mm^3$) | | Total brain volume ($mm^3$) | |
|---|---|---|---|---|---|---|---|---|
| | **β (95% CI) [a]** | **p value** | **β (95% CI) [a]** | **p value** | **β (95% CI) [a]** | **p value** | **β (95% CI) [a]** | **p value** |
| AVR, per SD | 1359.9 (160.3, 2559.6) | **0.026** | 1400.5 (4.3, 2796.7) | **0.049** | 534.1 (-744.9, 1813.1) | 0.413 | 1934.6 (-247.9, 4117.1) | 0.082 |
| CRAE, per SD | 1953.0 (755.4, 3150.7) | **0.001** | 2332.7 (939.0, 3726.3) | **0.001** | 1447.4 (170.3, 2724.4) | **0.026** | 3780.0 (1602.0, 5957.9) | **0.001** |
| CRVE, per SD | 993.7 (-201.1, 2188.6) | 0.103 | 1321.8 (-68.5, 2712.1) | 0.062 | 904.5 (-368.8, 2177.8) | 0.164 | 2226.2 (53.4, 4399.0) | **0.045** |
| FDa, per SD | 4091.4 (2880.4, 5302.3) | **<0.001** | 4791.5 (3382.5, 6200.6) | **<0.001** | 2231.7 (935.7, 3527.6) | **0.001** | 7023.3 (4819.3, 9227.2) | **<0.001** |
| FDv, per SD | 3135.1 (1934.5, 4335.8) | **<0.001** | 3699.4 (2302.4, 5096.4) | **<0.001** | 2086.0 (803.9, 3368.1) | **0.001** | 5785.4 (3601.9, 7968.9) | **<0.001** |
| Tortuosity of artery, per SD | 1140.7 (-56.6, 2338.0) | 0.062 | 1052.5 (-341.1, 2446.1) | 0.139 | 725.3 (-550.9, 2001.5) | 0.265 | 1777.8 (-400.1, 3955.8) | 0.110 |
| Tortuosity of vein, per SD | 1248.9 (55.8, 2442.0) | **0.040** | 1078.3 (-310.4, 2467.1) | 0.128 | 1414.8 (143.6, 2686.0) | **0.029** | 2493.2 (323.4, 4663.0) | **<0.001** |

[a] Adjusted for the actual CAIDE score, and brain MRI measuring positions.

Abbreviations: AVR: Arteriovenous ratio; CRAE: Central retinal arterial equivalent; CRVE: Central retinal venular equivalent; FDa: Fractal dimension artery; FDv: Fractal dimension vein.

**Supplementary Table 9: Associations between retinal vascular network parameters with dementia incidence**

| Retinal vascular parameters | All-cause dementia | | | Alzheimer's disease | | | Vascular dementia | | |
|---|---|---|---|---|---|---|---|---|---|
| | Incident cases | No.total | HR[a] (95% CI) | Incident cases | No.total | HR[a] (95% CI) | Incident cases | No.total | HR[a] (95% CI) |
| AVR | | | | | | | | | |
| Q1 | 140 | 8312 | Reference | 61 | 8312 | Reference | 28 | 8312 | Reference |
| Q2 | 78 | 6485 | **0.742 (0.562, 0.979)** | 36 | 6485 | 0.787 (0.521, 1.188) | 11 | 6485 | 0.531 (0.264, 1.067) |
| Q3 | 120 | 9280 | 0.832 (0.652, 1.062) | 61 | 9280 | 0.971 (0.680, 1.385) | 23 | 9280 | 0.822 (0.473, 1.428) |
| Q4 | 108 | 7259 | 1.024 (0.796, 1.317) | 45 | 7259 | 0.979 (0.665, 1.44) | 12 | 7259 | 0.599 (0.304, 1.181) |
| Q5 | 73 | 7527 | **0.676 (0.509, 0.898)** | 38 | 7527 | 0.807 (0.538, 1.212) | 6 | 7527 | **0.294 (0.121, 0.710)** |
| *P* for trend | | | 0.125 | | | 0.594 | | | **0.012** |
| Standardized continuous | | | **0.917 (0.842, 1.000)** | | | 0.972 (0.857, 1.103) | | | 0.715 (0.574, 0.890) |
| CRAE | | | | | | | | | |
| Q1 | 124 | 7866 | Reference | 59 | 7866 | Reference | 20 | 7866 | Reference |
| Q2 | 96 | 8013 | 0.811 (0.621, 1.058) | 45 | 8013 | 0.798 (0.541, 1.176) | 16 | 8013 | 0.855 (0.443, 1.651) |
| Q3 | 100 | 7328 | 0.926 (0.712, 1.206) | 47 | 7328 | 0.915 (0.623, 1.342) | 9 | 7328 | 0.527 (0.240, 1.157) |
| Q4 | 105 | 7515 | 0.985 (0.759, 1.278) | 47 | 7515 | 0.925 (0.63, 1.358) | 19 | 7515 | 1.139 (0.608, 2.136) |

| | | | | | | | | | |
|---|---|---|---|---|---|---|---|---|---|
| Q5 | 94 | 8141 | 0.830 (0.634, 1.085) | 43 | 8141 | 0.797 (0.538, 1.181) | 16 | 8141 | 0.910 (0.471, 1.758) |
| *P* for trend | | | 0.528 | | | 0.471 | | | 0.931 |
| Standardized continuous | | | 0.964 (0.883, 1.052) | | | 0.932 (0.819, 1.06) | | | 0.959 (0.768, 1.199) |
| CRVE | | | | | | | | | |
| Q1 | 87 | 7554 | Reference | 44 | 7554 | Reference | 10 | 7554 | Reference |
| Q2 | 127 | 8363 | 1.298 (0.988, 1.705) | 60 | 8363 | 1.210 (0.82, 1.786) | 15 | 8363 | 1.315 (0.591, 2.927) |
| Q3 | 92 | 7312 | 1.056 (0.788, 1.416) | 45 | 7312 | 1.021 (0.674, 1.547) | 14 | 7312 | 1.371 (0.609, 3.087) |
| Q4 | 101 | 7613 | 1.120 (0.841, 1.492) | 43 | 7613 | 0.943 (0.619, 1.435) | 22 | 7613 | 2.086 (0.988, 4.406) |
| Q5 | 112 | 8021 | 1.136 (0.858, 1.504) | 49 | 8021 | 0.982 (0.654, 1.477) | 19 | 8021 | 1.622 (0.754, 3.492) |
| *P* for trend | | | 0.840 | | | 0.969 | | | 0.097 |
| Standardized continuous | | | 1.003 (0.921, 1.093) | | | 0.950 (0.837, 1.077) | | | 1.152 (0.927, 1.432) |
| FDa | | | | | | | | | |
| Q1 | 168 | 7764 | Reference | 75 | 7764 | Reference | 28 | 7764 | Reference |
| Q2 | 126 | 7732 | **0.779 (0.618, 0.981)** | 67 | 7732 | 0.927 (0.667, 1.289) | 22 | 7732 | 0.829 (0.474, 1.450) |
| Q3 | 107 | 7798 | **0.686 (0.538, 0.874)** | 53 | 7798 | 0.759 (0.534, 1.080) | 11 | 7798 | **0.440 (0.219, 0.885)** |
| Q4 | 79 | 7820 | **0.548 (0.418, 0.716)** | 27 | 7820 | **0.417 (0.268, 0.649)** | 12 | 7820 | 0.539 (0.273, 1.063) |
| Q5 | 39 | 7749 | **0.332 (0.233, 0.473)** | 19 | 7749 | **0.359 (0.215, 0.599)** | 7 | 7749 | **0.421 (0.182, 0.976)** |

| | | | | | | | | | |
|---|---|---|---|---|---|---|---|---|---|
| *P* for trend | | | **<0.001** | | | **<0.001** | | | **0.007** |
| Standardized continuous | | | **0.790 (0.730, 0.855)** | | | **0.787 (0.701, 0.883)** | | | **0.778 (0.637, 0.950)** |
| FDv | | | | | | | | | |
| Q1 | 154 | 7789 | Reference | 79 | 7789 | Reference | 23 | 7789 | Reference |
| Q2 | 134 | 7827 | 0.897 (0.711, 1.130) | 64 | 7827 | 0.834 (0.599, 1.159) | 19 | 7827 | 0.869 (0.473, 1.596) |
| Q3 | 110 | 7766 | **0.772 (0.604, 0.986)** | 53 | 7766 | 0.722 (0.510, 1.024) | 13 | 7766 | 0.633 (0.320, 1.249) |
| Q4 | 71 | 7712 | **0.537 (0.405, 0.712)** | 28 | 7712 | **0.410 (0.266, 0.632)** | 15 | 7712 | 0.809 (0.421, 1.554) |
| Q5 | 50 | 7769 | **0.418 (0.303, 0.576)** | 17 | 7769 | **0.274 (0.162, 0.465)** | 10 | 7769 | 0.623 (0.295, 1.316) |
| *P* for trend | | | **<0.001** | | | **<0.001** | | | 0.197 |
| Standardized continuous | | | **0.793 (0.734, 0.857)** | | | **0.742 (0.665, 0.829)** | | | 0.825 (0.675, 1.008) |
| Tortuosity of artery | | | | | | | | | |
| Q1 | 124 | 7772 | Reference | 66 | 7772 | Reference | 14 | 7772 | Reference |
| Q2 | 120 | 7773 | 1.024 (0.797, 1.317) | 53 | 7773 | 0.849 (0.591, 1.219) | 21 | 7773 | 1.636 (0.832, 3.219) |
| Q3 | 101 | 7773 | 0.874 (0.672, 1.136) | 45 | 7773 | 0.730 (0.500, 1.067) | 14 | 7773 | 1.107 (0.528, 2.323) |
| Q4 | 91 | 7773 | 0.814 (0.621, 1.068) | 36 | 7773 | **0.603 (0.402, 0.906)** | 15 | 7773 | 1.243 (0.600, 2.578) |
| Q5 | 83 | 7772 | 0.768 (0.582, 1.015) | 41 | 7772 | 0.711 (0.481, 1.051) | 16 | 7772 | 1.393 (0.679, 2.859) |
| *P* for trend | | | **0.017** | | | **0.019** | | | 0.675 |

| | | | | | | | | | |
|---|---|---|---|---|---|---|---|---|---|
| Standardized continuous | | | 0.950 (0.864, 1.044) | | | 0.930 (0.807, 1.072) | | | 1.137 (0.938, 1.379) |
| Tortuosity of vein | | | | | | | | | |
| Q1 | 120 | 7772 | Reference | 60 | 7772 | Reference | 15 | 7772 | Reference |
| Q2 | 108 | 7773 | 0.959 (0.739, 1.244) | 49 | 7773 | 0.868 (0.595, 1.266) | 16 | 7773 | 1.158 (0.572, 2.342) |
| Q3 | 85 | 7773 | **0.755 (0.572, 0.997)** | 40 | 7773 | 0.710 (0.476, 1.060) | 12 | 7773 | 0.872 (0.408, 1.864) |
| Q4 | 88 | 7773 | 0.781 (0.593, 1.029) | 45 | 7773 | 0.798 (0.542, 1.175) | 11 | 7773 | 0.802 (0.368, 1.746) |
| Q5 | 118 | 7772 | 1.011 (0.784, 1.304) | 47 | 7772 | 0.804 (0.549, 1.177) | 26 | 7772 | 1.801 (0.954, 3.400) |
| *P* for trend | | | 0.581 | | | 0.217 | | | 0.152 |
| Standardized continuous | | | 1.054 (0.984, 1.129) | | | 0.987 (0.867, 1.124) | | | **1.133 (1.039, 1.236)** |

[a] Adjusted for the actual CAIDE score.

Abbreviations: AVR: Arteriovenous ratio; CRAE: Central retinal arterial equivalent; CRVE: Central retinal venular equivalent; FDa: Fractal dimension artery; FDv: Fractal dimension vein.

**Supplementary Table 10: The predictive performance of the algorithm-estimated CAIDE model integrating retinal vascular parameters for 14-year dementia risk via fundus images and compared to the actual CAIDE model**

| Metrics | Overall | Female | Male |
|---|---|---|---|
| **All-cause dementia** | | | |
| Events/Total | 509/35838 | 236/19393 | 273/16445 |
| AUC (Algorithm-estimated CAIDE model) | 0.683 (0.663-0.703) | 0.690 (0.661-0.720） | 0.667 (0.639-0.695） |
| AUC (+Retinal vascular parameters) | 0.721 (0.702-0.741) | 0.726 (0.697-0.755) | 0.735 (0.710-0.760) |
| DeLong's test p value for AUC comparisons | **0.002** | 0.059 | **<0.001** |
| **Alzheimer's disease** | | | |
| Events/Total | 236/35838 | 117/19393 | 119/16445 |
| AUC (Algorithm-estimated CAIDE model) | 0.679 (0.650-0.707) | 0.680 (0.640-0.721) | 0.674 (0.635-0.713) |
| AUC (+Retinal vascular parameters) | 0.730 (0.702-0.758) | 0.747 (0.705-0.789) | 0.754 (0.717-0.791) |
| DeLong's test p value for AUC comparisons | **0.005** | **0.014** | **0.001** |
| **Vascular dementia** | | | |
| Events/Total | 79/35838 | 35/19393 | 44/16445 |
| AUC (Algorithm-estimated CAIDE model) | 0.751 (0.710-0.792) | 0.759 (0.694-0.824) | 0.735 (0.681-0.790) |

| | | | |
|---|---|---|---|
| AUC (+Retinal vascular parameters) | 0.778 (0.734-0.822) | 0.847 (0.797-0.896) | 0.811 (0.750-0.872) |
| DeLong’s test p value for AUC comparisons | 0.353 | **0.012** | 0.095 |
